# Cardiovascular risk estimation and statin adherence *An historical cohort study*

**DOI:** 10.1101/2025.06.27.25330450

**Authors:** Samuel Finnikin, Brian Willis, Tim Evans, Brian Finney, Kwan Nok Chris Hui, Rani Khatib, Tom Marshall

## Abstract

**Background:** Adherence to statins for the primary prevention of cardiovascular disease (CVD) is low. There is evidence that some facets of the initiation consultation, or the initiating clinician, are associated with adherence. CVD risk estimation is fundamental to statin initiation and shared decision making (SDM), because the benefits of statins are proportional to CVD risk. Absence of a recorded CVD risk score before statin initiation therefore indicates SDM is unlikely. This study investigates whether, in primary prevention, SDM, using CVD risk score as a proxy measure, is associated with adherence to statins and CVD outcomes.

**Method:** A cohort of statin naïve patients aged 40-84 years initiated on statins for primary prevention between 2017 and 2020 was identified and categorised by the presence or absence of a CVD risk score at statin initiation. Statin adherence and persistence was determined from subsequent statin prescriptions. Multivariable modelling, accounting for potential confounders determined the association between a recorded CVD risk score and statin adherence and persistence. A secondary analysis investigated the relationship with subsequent CVD outcomes and death.

**Results:** 255,730 patients were included with a mean follow up of 4.6 years. 67.7% had a CVD risk score coded. The presence of a CVD risk score increased odds of adherence at the end of year one by 7% and reduced the chances of discontinuation by 8%. The composite outcome of CVD and all-cause mortality was decreased by 21% when a CVD risk score was present.

**Conclusion:** When a statin is initiated in the presence of a CVD risk score there is a small, but significant improvement in both adherence and persistence which could indicate that the quality of initiation consultations, and a focus on SDM, improves the utility of statins. Additionally, CVD risk scoring is associated with a large decrease in CVD/death which cannot fully be explained by improvements in statin adherence and persistence; an important finding necessitating further investigation.

**Clinical Perspective:** *What is new?:*

- We present a comprehensive assessment of the adherence to and persistence with statins for the primary prevention of CVD which, for the first time, is linked to the presence or absence of CVD risk scoring.
- CVD risk scoring is associated with improvements and both adherence and persistence as well as significant reductions in CVD and all-cause mortality.

*What are the clinical implications?:*

- There should be renewed focus on the content of statin initiation consultations ensuring that CVD risk is communicated using a shared decision-making approach.
- Further consideration should be given to the important discrepancy in CVD and death seen when CVD risk scoring is, and is not, used in practice.

## Introduction

Optimal use of lipid lowering therapies (LLTs) is one of the cornerstones of cardiovascular disease (CVD) prevention strategies worldwide and, as a result, statins, the most used class of LLTs, are one of the most commonly prescribed class of medications. (1) However, adherence to statins is known to be low, with roughly half of people being non-adherent after a year. (2-4) Poor adherence to statins used for primary prevention is associated with an increased risk of cardiovascular events. (5)

Statins for the primary prevention of CVD are usually recommended to patients based on an estimate of their overall risk of CVD. (6-8) This strategy is adopted because the benefit a patient may gain from a statin is proportional to their CVD risk, whilst the disadvantages and costs of the medication are, essentially, unrelated to the underlying CVD risk. Therefore, at low CVD risk, prescribing statins may not be cost effective and/or the net benefit may not be sufficient at an individual level based on the patient’s preferences and values. (9) Thus, CVD risk estimation is a necessary component of the offer and acceptance of a statin and should be discussed with patients using a Shared Decision Making (SDM) approach. Shared decision making is the accepted and preferred approach to medical decision making consisting of a thorough discussion of the options available to a patient (including doing nothing), along with their risks and benefits, before agreeing on the option that is right for the individual. (10) It follows that CVD risk estimation is a necessary component of statin initiation decisions as a fully informed discussion about the benefits and risks of statins is not possible without this information.

In the UK, QRISK is used for CVD risk estimation. (11) Several iterations of this algorithm have been used, with QRISK3 being the most current one used in practice. Previous studies using UK primary care data have shown that many patients are initiated on statins without a QRISK score recorded in the patient’s record. (12) Moreover, when QRISK is not recorded, the principal predictor of the decision to initiate statins is the total cholesterol level whereas, when it is recorded, the CVD risk estimate is the principal predictor. (13) The presence or absence of a record of a QRISK score is, therefore, an indicator of different decision-making processes. If a QRISK score is not coded in the patients record when statins are initiated, the evidence suggests CVD risk estimates are not discussed at the time of statin initiation and SDM is less likely to have taken place. Conversely, a QRISK score coded at the time of statin initiation indicates that the necessary information required for SDM was available to the clinician during the consultation. Thus, it is reasonable to hypothesise that there is an association between the coding of a CVD risk estimate and SDM when statins are initiated for the primary prevention of CVD.

It has been argued that adherence to medications could be improved through better SDM at initiation, (14-18) and there is evidence that some facets of the initiation consultation, or the clinician doing the initiation, are associated with adherence. (19, 20) Establishing a link between SDM and adherence is, however, not straightforward; largely because measuring SDM in practice is difficult to do. (21) However, having established that the presence or absence of a QRISK score is associated with a different initiation decision process, and that CVD risk estimation is necessary for SDM, there is an opportunity to explore the link between SDM and adherence in a new way.

The objective of this study is to investigate whether evidence of SDM (as determined by the recording of QRISK code where indicated) at the point of statin initiation is associated with adherence to, and persistence with, statins and CVD outcomes.

## Method

This cohort study used CPRD Aurum (Version 2023.12.001), and the study design and data extraction were facilitated using the Dexter software. (22, 23) All statin naïve patients aged 40-84 years (inclusive) initiated on statins for primary prevention between 2017 and 2020 were included. To only include patients where a QRISK calculation was appropriate, patients were excluded if they had existing CVD (or a CVD event within 60 days of statin initiation to account for any coding delay), Type 1 diabetes mellitus, chronic kidney disease (CKD) stages 3-5 or familial hypercholesterolaemia (either coded or presumed due to total cholesterol being ≥9.0 mmol/l). The population was split into two cohorts; those with a coded QRISK score and those without. A QRISK score coded within the 60 days prior to statin initiation was regarded as being associated with the statin initiation. This time period was chosen because it was considered clinically plausible that a CVD risk estimation within 60 days is likely to have been calculated because of a risk factor update (e.g. new lipid profile result) and resulted in the statin initiation consultation. This QRISK would therefore be logically linked to the initiation decision.

The index date was the date of the first statin prescription. Patients were eligible for inclusion from the earliest of the following dates: study start date, registration date plus 1 year, and age 40; until the earliest of the following dates: age 85, study end date, CVD diagnosis, or other excluding diagnosis. Patients were excluded if they left the database within 2 years of the index date as a minimum of 2 year follow up was required. Patients were followed up until the earliest of the following dates: study end period, exit from the database, a new CVD diagnosis or death.

Baseline characteristics and variables that may be associated with adherence were identified along with coded QRISK scores (latest recorded in the period 60d prior to initiation up until statin initiation) and statin type and dose. Relevant comorbidities were hypertension, atrial fibrillation, type 2 diabetes mellitus, inflammatory arthritis and severe enduring mental illness. These were combined into a single ordinal variable. QRISK2 and QRISK3 scores were both extracted and treated in the same way and therefore not referred to separately in this manuscript. All subsequent statin and other LLT prescriptions (ezetimibe, bempedoic acid and fibrates) following initiation were identified (type, quantity and duration of prescription in days). Lipid profiles and new CVD diagnoses were the other outcome variables.

### Statistics

The primary outcome of adherence was measured using the Medication Possession Ratio (MPR). (24, 25) The quantity of drug prescribed, rather than the ‘days’ prescribed, was used for the prescription duration as it was felt that this would provide more accurate data. Statins (and other LLTs) are prescribed as a once daily dosing regimen but the number of ‘days’ entered by the clinician can be inconsistent with this. A sensitivity analysis was undertaken only including patients where number of days is equal to the quantity of drug prescribed to check this assumption. Patients were excluded from adherence analysis if they were only issued one prescription of a statin (but these will be included in persistence analysis). Patients were be classified as being adherent if their MPR was greater than 0.8 in a 12 month period as this threshold is consistently used for research purposes. (26) Adherence (MPR) was be calculated for each completed 12-month period of follow-up to assess for trends.

For persistence, patients were identified as having discontinued statin therapy if there was no new prescription 180 days after the expected end of their supply. There is no consensus on the permissible gap (27) in persistence research but 180 days is commensurate with other research (4) and considered clinically appropriate by the authors with experience of English prescribing practices. Persistence was defined as the time between initiation and the final prescription plus the duration of the last prescription. (26) A sensitivity analysis was performed including all oral LLTs prescribed following statin initiation.

Patients who restarted their medication after a period of discontinuation >180 days were identified but prescriptions after a period of discontinuation were not included in adherence and persistence analysis based on the assumption that further discussion about the medication may have influenced the decision to recommence so the link with the initiation consultation is less robust.

Multivariable logistic regression modelling was used to establish the impact of variables on adherence in the first 12 months (as measured by MPR) with presence or absence of QRISK score being the main variable of interest and practice ID as a random effect (change from protocol as prescriber level data was not available.) Mediator analysis was undertaken using Med4Way (28) to establish whether the association between QRISK coding and CVD outcomes and death were mediated by adherence and persistence.

Two time-to-event analyses using Cox regression for the outcomes of statin discontinuation and CVD outcomes and death were performed. In both cases, QRISK coding was the main predictor variable. Statistical significance for all models was adjusted for multiple testing using the Bonferroni technique. (29)

As prescribing data is electronic and recorded automatically it is assumed that the absence of a prescription means that none was issued. If QRISK score is missing it will be presumed to be absent. Previous research has shown that when QRISK2 is not recorded it does not seem to influence prescribing decisions which provides validity to this assumption. (13) Categories for missing data were used for categorical variables. All analysis was performed using StataSE V18.

## Results

The cohort comprised 255,752 patients from 1771 practices (see strobe diagram figure 1.) The characteristics of the included patients are shown in table 1 split by the presence or absence of a QRISK score (within 60 days of statin initiation). Two thirds of patients had a QRISK score coded (173,051, 67.7% 95% confidence interval (CI) 67.5-67.8%) and the groups were statistically different by all measures apart from the incidence of raised liver transaminases and the combined variable of CVD diagnoses or death (p value for significance adjusted to 0.0036 by Bonferroni correction). Notably, those patients with a QRISK score, when compared to those without, were more likely to be male (54.3% Vs 49.2%), older (mean age 63.3% Vs 59.6%), from areas of lower deprivation (16.7% in the 1^st^ IMD quintile compared with 10.3%), and have White ethnicity (85.1% Vs 79.1%). The relationship between QRISK coding and demographic variables is expanded upon through logistic regression in supplementary table S1. Atorvastatin was the most common statin prescribed making up 93.4% initial prescriptions, and 89.7% of all prescriptions (supplementary table S2).

**Table 1:**
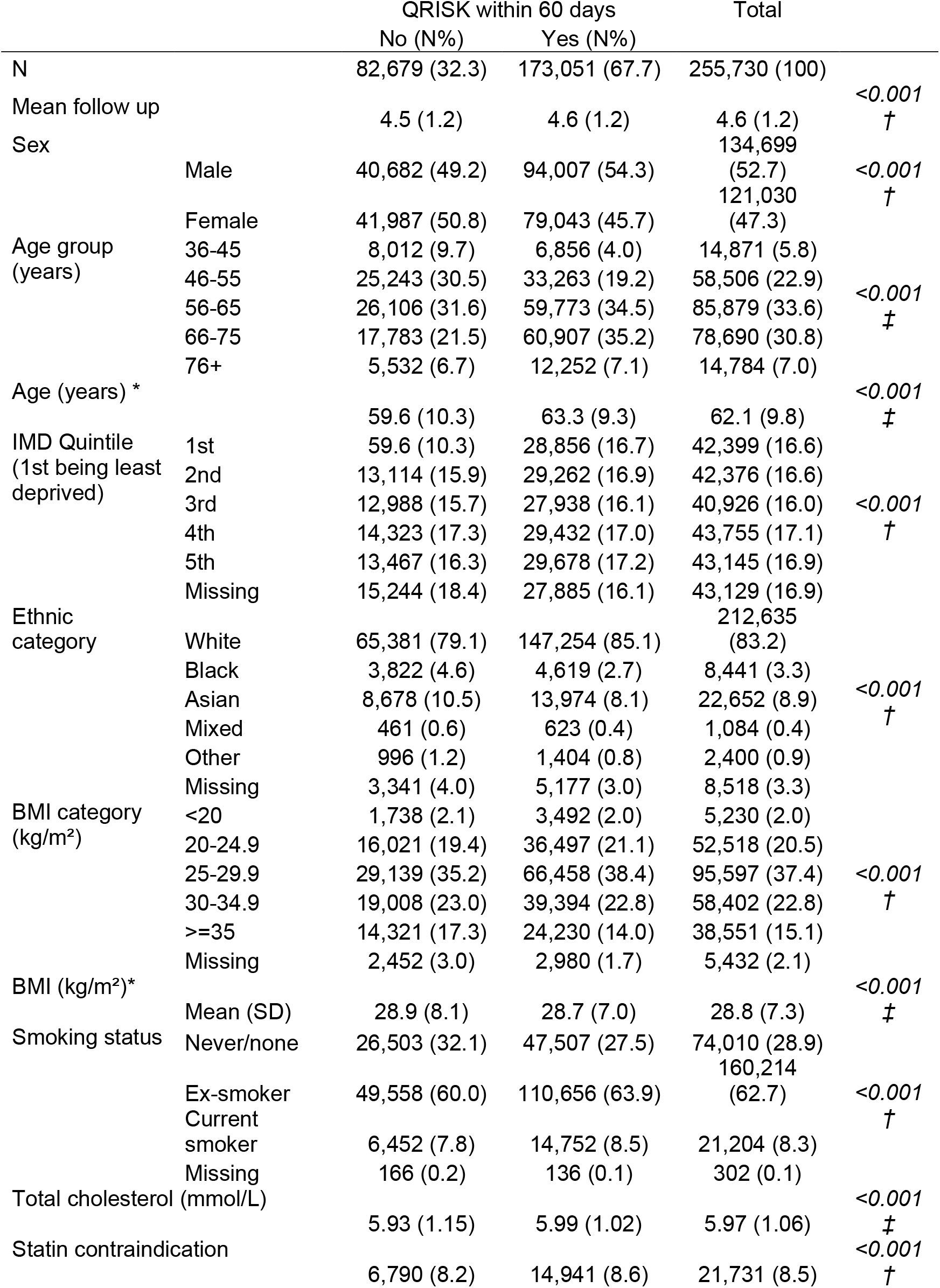
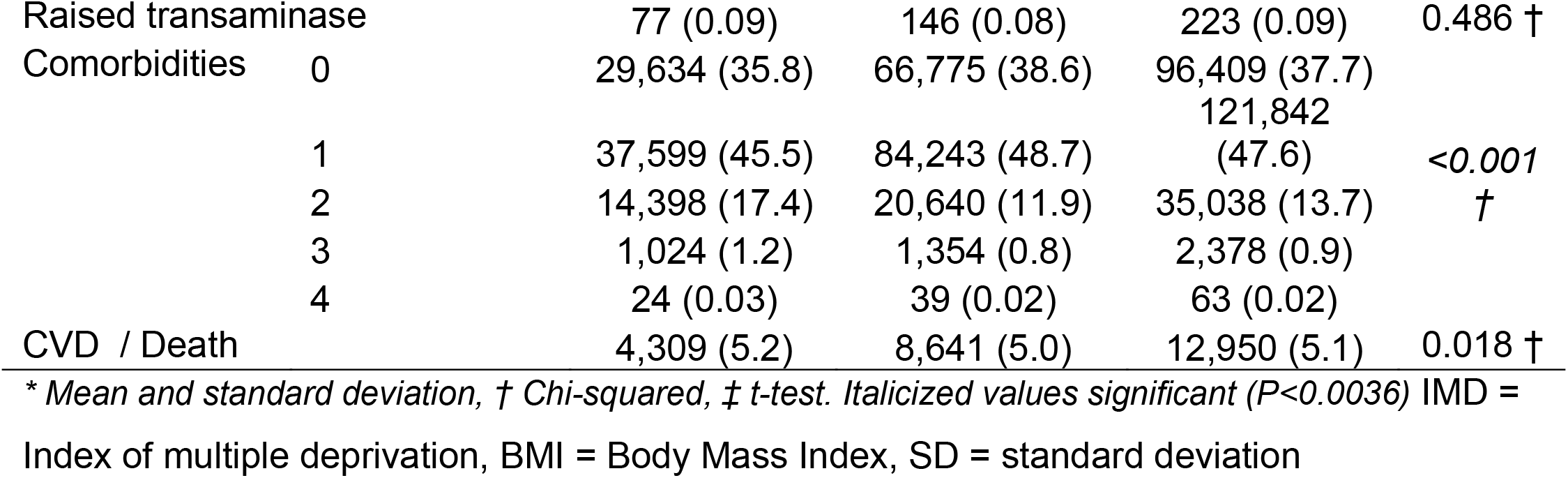
Cohort characteristics.

**Table 2:**
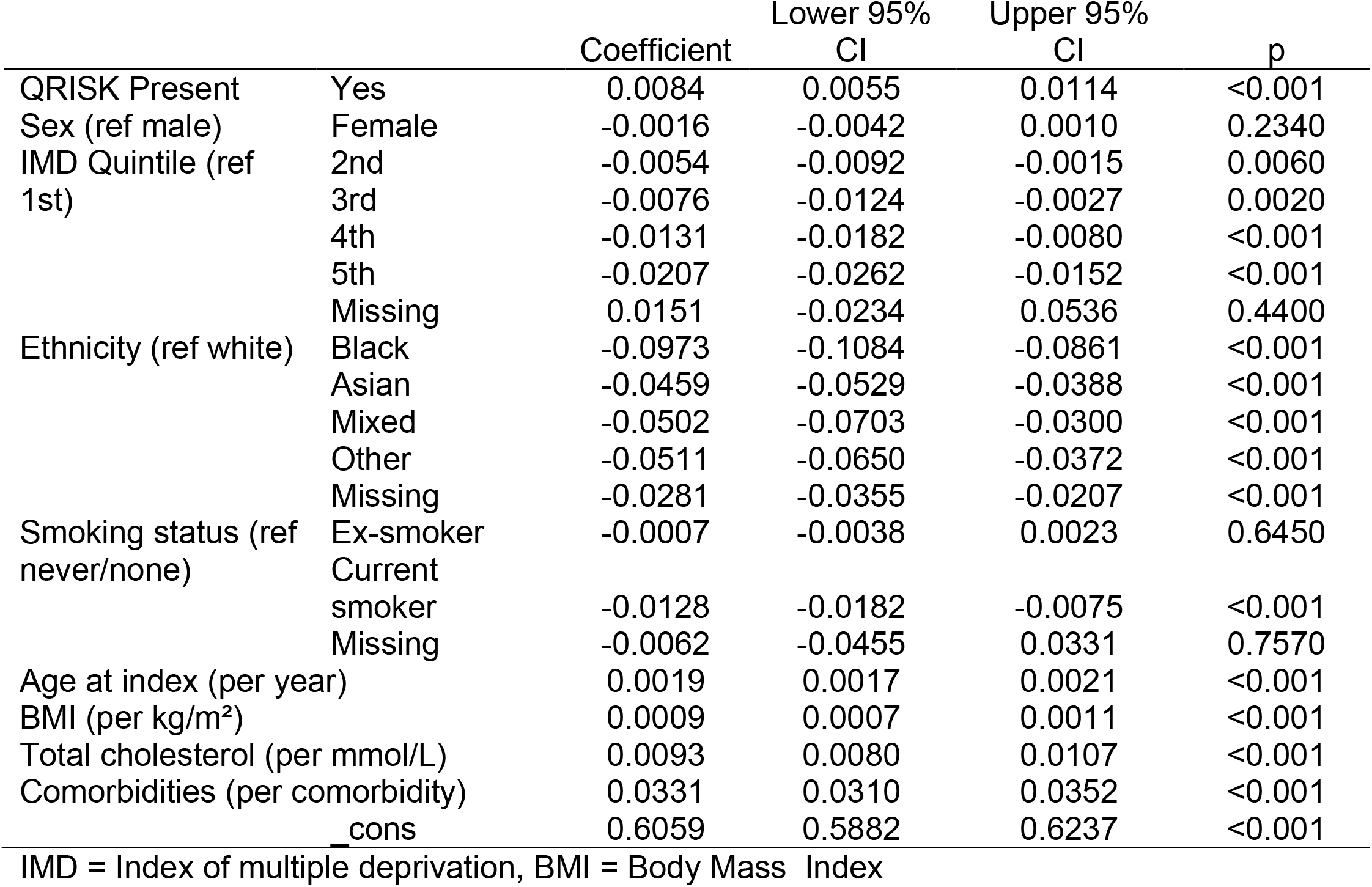
Effects on variables on MPR in year 1 (linear regression)

**Figure 1.**
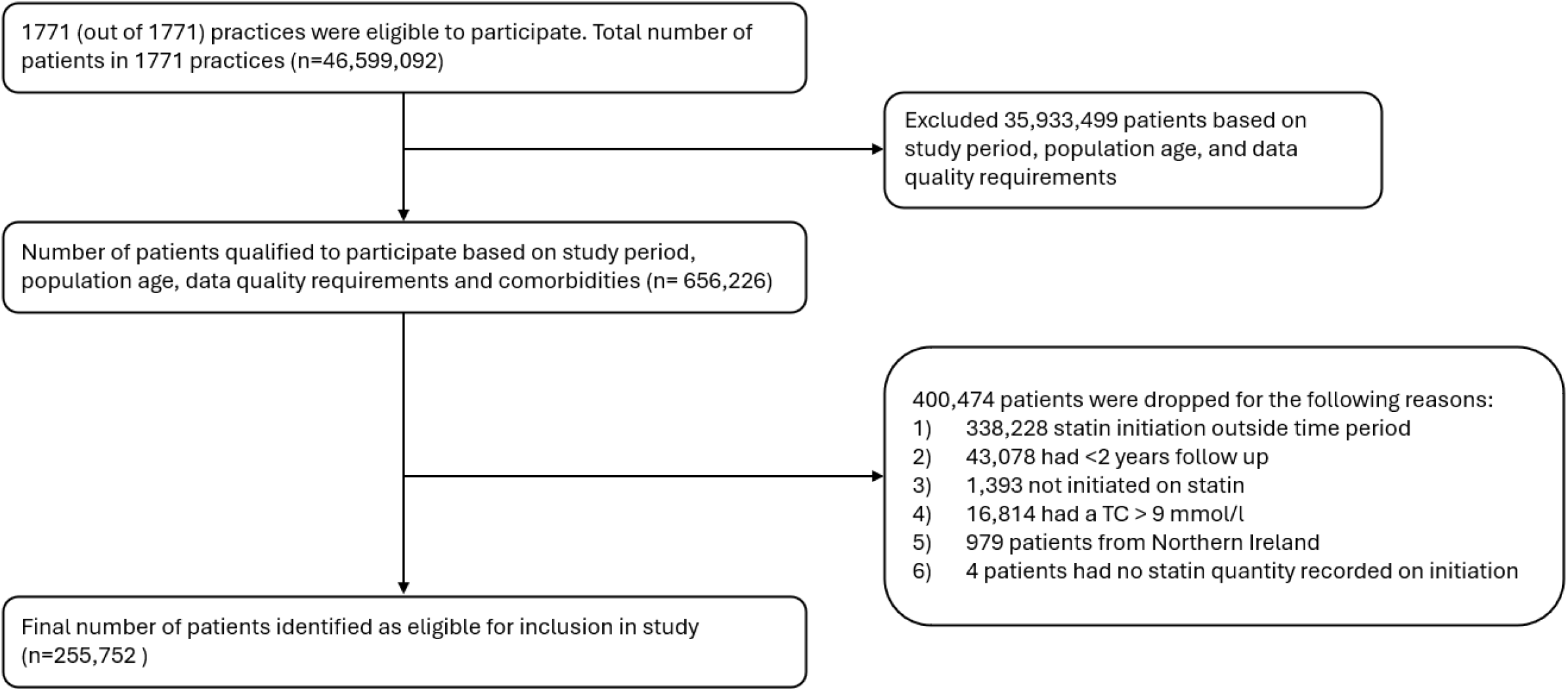
STROBE diagram

The MPR in the first year of follow up (excluding people who only received the index prescription) was 82.4% (95%CI 82.3-82.6%) in patients with a QRISK score compared with 80.9% (95%CI 81.8-82.1%) in those without a QRISK score (p<0.001). Figure 2 shows how the trend in adherence (MPR>0.8) changed over the duration of observation and is consistently around 3% higher for people with a QRISK score coded compared to those without. Linear regression modelling using MPR in year 1 as a continuous variable shows that the presence of a QRISK score is associated with increased adherence, alongside increasing age, BMI, baseline total cholesterol and the number of comorbidities (table 1). Female sex, increasing deprivation and non-white ethnicities are negatively associated with MPR with very little collinearity between variables (mean variance inflation factor 1.21).

**Figure 2.**
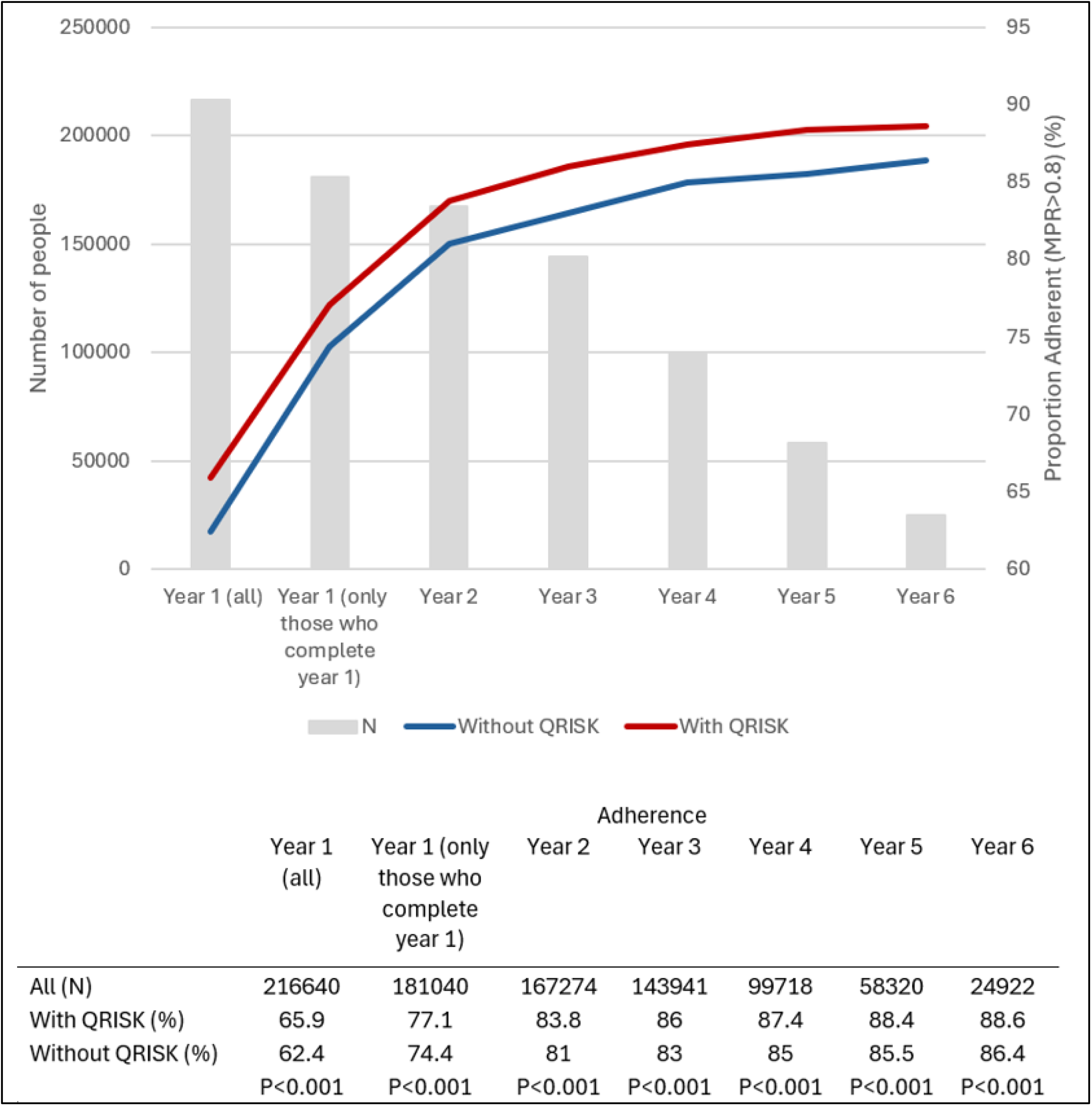
Number of participants per year of follow up and adherence by QRISK status

When adherence is modelled as a binary variable (MPR>0.8) using logistic regression (supplementary table S2), the presence of QRISK increases the odds of adherence by 7% (OR 1.07. 95% CI 1.05-1.10). The presence of co-morbidities has the highest contribution to adherence with a 26% increase in adherence per co-morbidity and all non-white ethnicities have significantly lower adherence ranging from 61% lower (Black ethnicity) to 37% lower (mixed ethnicity).

Overall, 100,127 (39.2%, 95%CI 39.0-39.3%) people discontinued their statin for >180 days during the observation period, 75,280 (29.4%, 95%CI 29.3-29.6%) in the first year. The median time to discontinuation was 110 days (IQR 56-369.) Of those who discontinued, 51,920 (51.9%) restarted a statin or other LLT with a median discontinuation period of 458 days (IQR 306-864 days). Supplementary table S3 describes which medications are issued before and after discontinuation and supplementary table S4 shows the number of discontinuations per year split by QRISK status. Figure 3 shows the differential discontinuation of statins for people with or without QRISK in an unadjusted Kaplan-Meier survival analysis. The results of the Cox regression analysis (adjusted for confounders and clustering at practice level) are shown in table 3. Presence of QRISK coding reduced discontinuation by 7% (HR 0.93, 95% CI 0.91-0.94) and the risk of discontinuation was reduced by 9% for each 1 mmol/l increase in total cholesterol and 25% for each additional comorbidity. Non-white ethnicity and higher levels of deprivation were associated with an increased hazard for discontinuation.

**Table 3:**
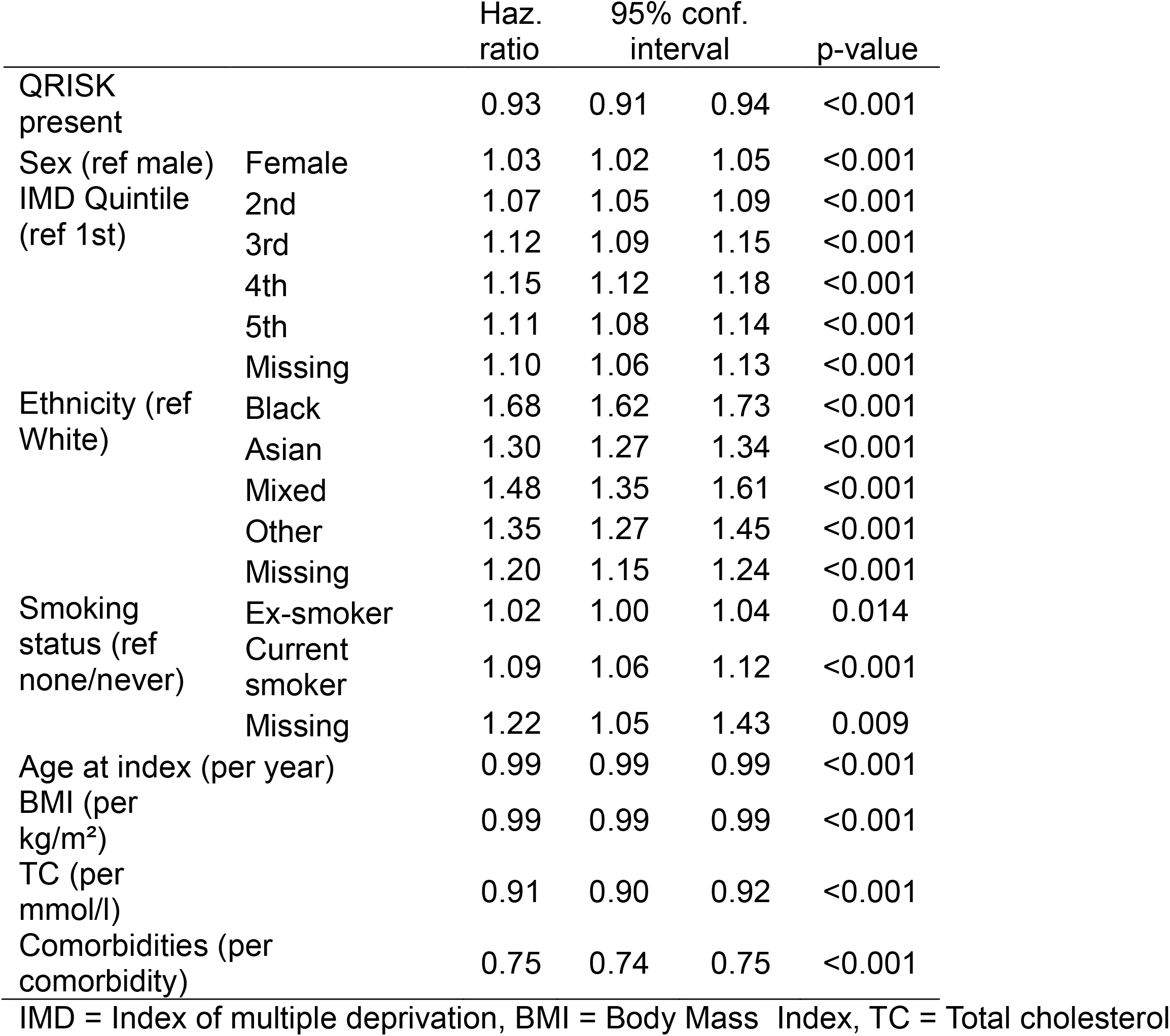
Adjusted Cox proportional hazards model for discontinuation.

**Figure 3.**
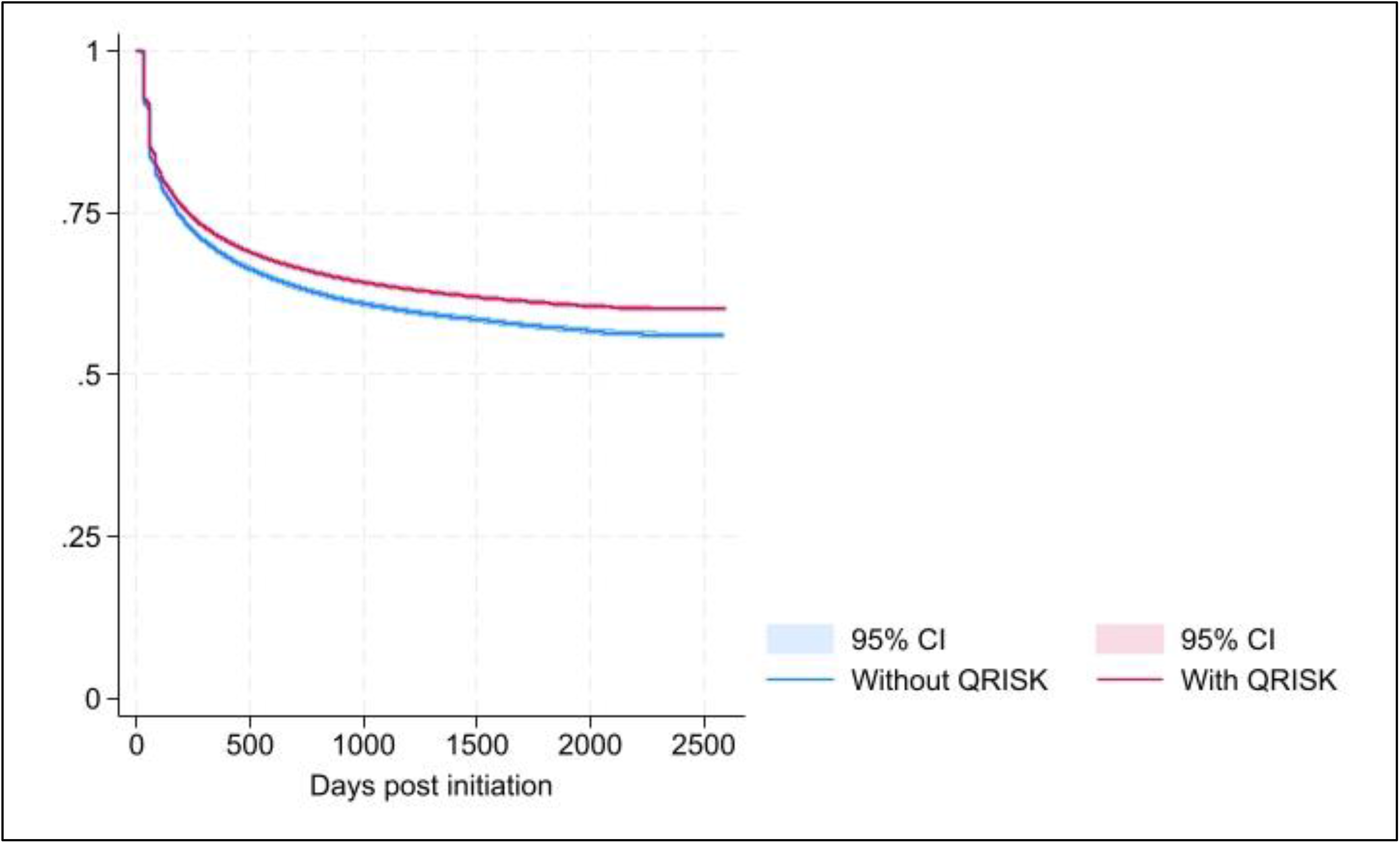
Kaplan-Meier curve showing cumulative discontinuation rate for patient with and without QRISK (unadjusted)

A total of 12,950 (5.1%, 95%CI 5.0-5.2%) patients died or had a new diagnosis of CVD during follow up, with similar proportions in both those with, and those without, a QRISK score. However, when adjusting for confounders in the Cox regression analysis, the presence of a QRISK score was associated with a 21% reduction in the risk of CVD/death (HR 0.79, 95%CI 0.76-0.82) (table 4). Current smoking was associated with the highest risk of CVD/death (2.02, 95%CI 1.89-2.16) and increasing risk was seen with increasing deprivation, but Black and Other ethnicities had lower hazard ratios when compared with White ethnicity (0.68 (95%CI 0.6-0.79) and 0.64 (95%CI 0.51-0.81) respectively.)

**Table 4:**
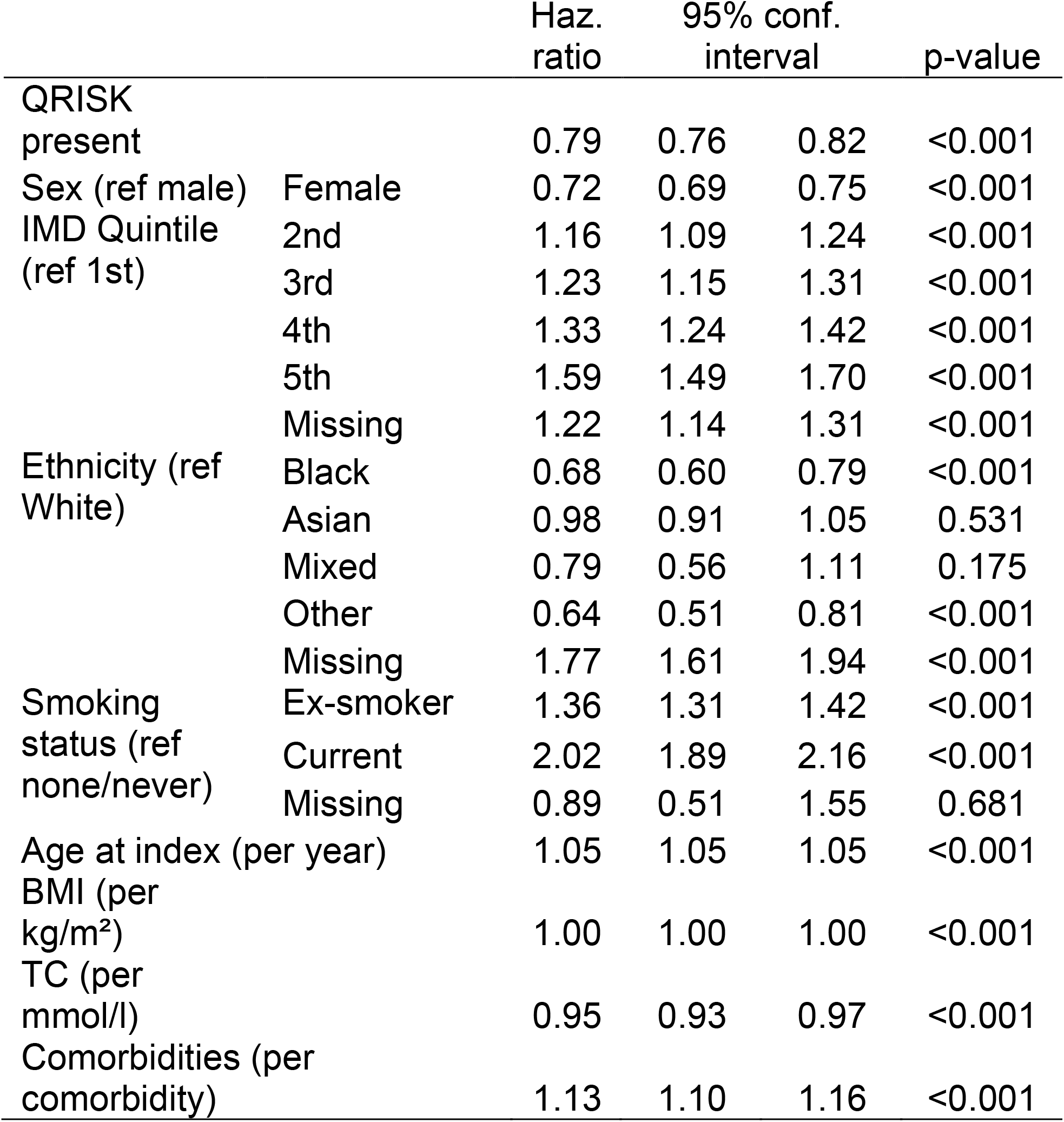
Adjusted Cox proportional hazards model for CVD/Death.

Mediation analysis showed there was a small but significant proportion of the reduction in CVD/death observed is mediated through improvement in adherence with little interaction between QRISK and MPR (supplementary table S5). Similarly, mediation analysis looking at time on treatment showed that QRISK scoring led to longer time on treatment which reduced the odds of CVD/death but most of the direct effect is not explained by treatment duration.

The two sensitivity analyses did not reveal any significant differences when adherence and persistence were examined for all LLTs or when only prescriptions where the number of days matched the quantity of tablets (supplementary table S6).

## Discussion

In this cohort, the presence of QRISK coding around the time of statin initiation is associated with modest but significant improvements in both adherence to, and persistence with, statins. QRISK coding is also associated with a significant (21%) reduction in the composite outcome of subsequent CVD diagnoses or all-cause mortality. It is implausible that this large reduction is fully explained by the combination of improved adherence and persistence even though these improvements are likely to have an additive effect on CVD outcomes. It is unlikely that the CVD risk in the cohort without QRISK coding is higher than the cohort with QRISK coding since age, sex and smoking status are the main predictors of CVD risk and all these variables are more favourable (in terms of CVD risk) in the uncoded cohort. This suggests that QRISK coding is associated with other factors that impact on CVD risk that are not captured in this study. It could be that when QRISK coding is used, there is a more holistic discussion about CVD risk which results in improvements in other risk factors (for example smoking cessation or better blood pressure control). This is plausible since the act of populating the QRISK variables focuses attention (from both the clinician and patient) on these other risk factors. Or it could simply be that QRISK coding is a proxy for a higher standard of clinical care which permeates the whole of an individual’s primary healthcare delivery resulting in better outcomes. Further research would be required to explore this important finding further.

### Comparisons with existing research

Two thirds of patient initiated on a statin had a QRISK score coded in their record in the 60 days prior to initiation. This is considerably higher than the 27.1% reported between 2012-15 but consistent with the increasing trend noted at that time, (30) and similar to the reported coding rate (within 6 months of initiation) in 2017/18. (31) QRISK recording was higher in males, older age groups and people living with lower deprivation. Part of these demographic differences may be explained by the differences in the demographics of people attending the NHS Health Check programme which focuses on CVD risk and sees higher uptake in low deprivation areas and amongst older people. (32)

The adherence rates observed in the first year (>80%) is higher than observed in most other studies in systematic reviews (3, 33) It is worth noting, however, that although an MPR of 0.8 is quite consistently used as a cutoff for adherence, there is variation in how MPR is calculated between studies and none of the included studies were UK based. In these reviews, increasing age, male sex and lower levels of deprivation have previously been found to be associated with improved adherence, and this is consistent with our findings.

Two studies examined ethnicity and adherence, both from the US, and both found that non-white ethnicities had lower rates of adherence. The impact of ethnicity on adherence was notably high, particularly in the Asian, mixed and other categories. A review into statin non-adherence also found that female sex, non-white ethnicities and low socioeconomic status were associated with worse adherence. (34) We found a strong association between adherence and increasing comorbidities. In the literature, the association between adherence and comorbidities is complicated by differences between different diagnoses as well as between primary and secondary prevention but some studies have also found that increasing comorbidities increase adherence in line with our findings. (3) Co-morbidities and co-prescribing are likely to be associated and, although we did not measure co-prescribing, there is similar conflicting evidence on the impact of polypharmacy on adherence in the preventive setting. (4, 35-38).

Existing literature on persistence is highly variable in the definition of persistence, whether reported as a dichotomous or continuous variable and in the duration of follow up meaning there are no direct comparisons with our findings. (33) Of the studies with longer durations of follow up there is large variation in persistence with an Israeli study reporting >75% of patients discontinuing statin therapy by 2 years; (39) an American Veteran cohort finding discontinuation rates of around 37% after 675 days (40) and a Danish cohort (mixed primary and secondary prevention) observing only 16% discontinuation at 3 years. (41) The large impact that co-morbidities have on persistence may be a result of increased opportunities to review medication that come with annual reviews of the long-term conditions that we included in our co-morbidities list. These review, incentivised by the Quality and Outcomes Framework (QOF) in England (42), are opportunities to reinforce medicine taking practices and address concerns. Frequent contact with physicians, particularly when it’s the same clinician who initiated the statin, is associated with improved adherence and persistence. (43)

Finally, there has been found to be a ‘clinicianclinician component’ to statin adherence. An American study showed that when prescriptions are written by a patient’s own primary care provider adherence rates are higher (OR 1.12), (44) and a questionnaire based study found that the type and level of information shared during consultations and the quality of consultation positively correlated with statin adherence. (45) A Danish study showed the ‘health management skills’ of a physician (defined as the ability to facilitate adherence with statins) positively impact health outcomes. (19) These suggest that the interpersonal skills and competencies of the clinician impact on subsequent statin adherence. It is likely that the association we have found between QRISK scoring and adherence, persistence and, ultimately, CVD outcomes and death is, at least in part, a proxy measure for the clinician component statin initiation consultations.

### Strengths and limitations

This study is a comprehensive examination of primary prevention statin adherence and persistence and, for the first time, harness the power of a large English primary care dataset to investigate whether some facet of the statin initiation consultation (as measured by QRISK coding) impacts on subsequent statin use and outcomes. By using electronic prescribing records, we have a complete understanding of prescriptions issued to patients in a dataset that is generalisable to the English population allowing a high degree of precision in the results. We also considered the potential impact of other LLTs in our analysis given increasing use of these for people intolerant to statins but found this did not significantly impact the results.

One key limitation of this type of study is that we are using prescriptions issued as a proxy for medications taken. It is acknowledged that, following prescribing, the medication must be dispensed, collected and, ultimately, taken by the patient and adherence/persistence could be affected at any point in this chain. Thus, the adherence/persistence levels we observed are the maximum possible levels, with potential for the real numbers to be reduced at several points. Other ways to measure adherence/persistence are possible, including directly observing medicine taking, using electronic pill dispensers or self-reporting of medicine taking. Each has its limitations, (4) and none would allow the large number of participants and long follow up we have achieved in this study. Given that statins are long term, repeat prescriptions, it is likely that the link between prescription and medicine taking is stronger than one-off or short-term prescriptions since the patient keeps coming back for a top-up of their supply indicating a likelihood of a reduced stock in the patient’s home, although some patients may stockpile medications.

Some patients restarted LLT after a period of discontinuation. The prescriptions after restarting were not included in the analysis and this may weaken the association with adherence/persistence and CVD outcomes as patients may be more adherent on restarting but the link with the initial statin consultation will have been lost.

Finally, we are assuming that when a QRISK score is coded in the record that it is communicated to the patient and used in the initiation consultation through a SDM process. This won’t always be the case and thus the association between QRISK scoring and adherence/persistence and outcomes would be diluted. It is also possible that clinicians engage in SDM guided by CVD risk estimates without a coded QRISK score but, as stated previously, this is not likely to be common given subsequent prescribing patterns observed. (13) Additionally, there is a possibility of residual confounding with a retrospective cohort study which may explain some of the large differences in CVD outcomes and death that we observed.

### Implications

There is a clear indication in our findings that the presence of a QRISK score at the time of statin initiation is associated with improvements in adherence and persistence. We argue that this is due to a better quality of consultation when a QRISK score is present and a higher likelihood that SDM has taken place. Thus, whilst there is understandable focus on ensuring the right people are offered a statin, we must also pay attention to *how* a statin is initiated to ensure that all patients have a high quality, risk informed discussion. Additionally, the large decrease in the development of CVD or all-cause mortality associated with the presence of QRISK coding cannot be explained by statin adherence/persistence alone and warrants further investigation. It could be that the use of QRISK is a marker of the quality of care that patients are receiving which could be potentially useful in the quality assurance processes of English primary care.

## Data Availability

Primary data is not available through the authors due to data sharing restrictions but is available to researchers through CPRD (subject to approvals). Code lists used in data extraction and processing are available on request from the corresponding author.

https://www.cprd.com/

## Data access and sharing

SF had full access to data used in this research. Primary data is not available through the authors due to data sharing restrictions but is available to researchers through CPRD (subject to approvals). Code lists used in data extraction and processing are available on request from the corresponding author.

### Acknowledgments

None

## References

1. Stewart J, Manmathan G, Wilkinson P. Primary prevention of cardiovascular disease: A review of contemporary guidance and literature. JRSM cardiovascular disease. 2017;6:2048004016687211.

2. Lemstra M, Blackburn D, Crawley A, Fung R. Proportion and risk indicators of nonadherence to statin therapy: a meta-analysis. Canadian Journal of Cardiology. 2012;28(5):574–80.

3. Hope HF, Binkley GM, Fenton S, Kitas GD, Verstappen SM, Symmons DP. Systematic review of the predictors of statin adherence for the primary prevention of cardiovascular disease. PLoS One. 2019;14(1):e0201196.

4. Ofori-Asenso R, Jakhu A, Zomer E, Curtis AJ, Korhonen MJ, Nelson M, Gambhir M, Tonkin A, Liew D, Zoungas S. Adherence and persistence among statin users aged 65 years and over: a systematic review and meta-analysis. The Journals of Gerontology: Series A. 2018;73(6):813–9.

5. Martin-Ruiz E, Olry-de-Labry-Lima A, Ocaña-Riola R, Epstein D. Systematic review of the effect of adherence to statin treatment on critical cardiovascular events and mortality in primary prevention. Journal of cardiovascular pharmacology and therapeutics. 2018;23(3):200–15.

6. Chou R, Cantor A, Dana T, Wagner J, Ahmed AY, Fu R, Ferencik M. Statin use for the primary prevention of cardiovascular disease in adults: updated evidence report and systematic review for the US Preventive Services Task Force. Jama. 2022;328(8):754–71.

7. World Health Organization. WHO package of essential noncommunicable (PEN) disease interventions for primary health care. 2020.

8. Mach F, Baigent C, Catapano AL, Koskinas KC, Casula M, Badimon L, Chapman MJ, De Backer GG, Delgado V, Ference BA. 2019 ESC/EAS Guidelines for the management of dyslipidaemias: lipid modification to reduce cardiovascular risk: The Task Force for the management of dyslipidaemias of the European Society of Cardiology (ESC) and European Atherosclerosis Society (EAS). European heart journal. 2020;41(1):111–88.

9. Albarqouni L, Doust J, Glasziou P. Patient preferences for cardiovascular preventive medication: a systematic review. Heart. 2017;103(20):1578–86.

10. NICE. Shared decision making. London: NICE; 2021.

11. Hippisley-Cox J, Coupland C, Brindle P. Development and validation of QRISK3 risk prediction algorithms to estimate future risk of cardiovascular disease: prospective cohort study. BMJ. 2017;357.

12. Finnikin S, Ryan R, Marshall T. Statin initiations and QRISK2 scoring in UK general practice: a THIN database study. Brit J Gen Pract. 2017;67(665):e881–e7.

13. Finnikin S, Willis BH, Ryan R, Evans T, Marshall T. Factors predicting statin prescribing for primary prevention: a historical cohort study. British Journal of General Practice. 2021;71(704):e219–e25.

14. Banning M. Older people and adherence with medication: a review of the literature. International journal of nursing studies. 2008;45(10):1550–61.

15. Bauer AM, Parker MM, Schillinger D, Katon W, Adler N, Adams AS, Moffet HH, Karter AJ. Associations between antidepressant adherence and shared decision-making, patient– provider trust, and communication among adults with diabetes: diabetes study of northern California (DISTANCE). Journal of general internal medicine. 2014;29:1139–47.

16. LeBlanc A, Herrin J, Williams MD, Inselman JW, Branda ME, Shah ND, Heim EM, Dick SR, Linzer M, Boehm DH. Shared decision making for antidepressants in primary care: a cluster randomized trial. JAMA internal medicine. 2015;175(11):1761–70.

17. Polinski JM, Kesselheim AS, Frolkis JP, Wescott P, Allen-Coleman C, Fischer MA. A matter of trust: patient barriers to primary medication adherence. Health Education Research. 2014;29(5):755–63.

18. Schoenthaler A, Rosenthal DM, Butler M, Jacobowitz L. Medication adherence improvement similar for shared decision-making preference or longer patient-provider relationship. The Journal of the American Board of Family Medicine. 2018;31(5):752–60.

19. Simeonova E, Skipper N, Thingholm PR. Physician health management skills and patient outcomes. J Hum Resour. 2022.

20. Kampouraki E, Wardman D, McFarlane L, Khatib R. 210 A positive consultation experience is associated with better statin adherence. BMJ Publishing Group Ltd and British Cardiovascular Society; 2023.

21. Barr PJ, Elwyn G. Measurement challenges in shared decision making: putting the ‘patient’in patient-reported measures. Health Expectations. 2016;19(5):993–1001.

22. Wolf A, Dedman D, Campbell J, Booth H, Lunn D, Chapman J, Myles P. Data resource profile: clinical practice research Datalink (CPRD) aurum. Int J Epidemiol. 2019;48(6):1740–g.

23. Gokhale KM, Chandan JS, Toulis K, Gkoutos G, Tino P, Nirantharakumar K. Data extraction for epidemiological research (DExtER): a novel tool for automated clinical epidemiology studies. European journal of epidemiology. 2021;36(2):165–78.

24. Andrade SE, Kahler KH, Frech F, Chan KA. Methods for evaluation of medication adherence and persistence using automated databases. Pharmacoepidemiology and drug safety. 2006;15(8):565–74.

25. Linden A. Assessing medication adherence using Stata. Stata J. 2019;19(4):820–31.

26. Cramer JA, Roy A, Burrell A, Fairchild CJ, Fuldeore MJ, Ollendorf DA, Wong PK. Medication compliance and persistence: terminology and definitions. Value Health. 2008;11(1):44–7.

27. Cramer JA, Roy A, Burrell A, Fairchild CJ, Fuldeore MJ, Ollendorf DA, Wong PK. Medication compliance and persistence: terminology and definitions. Value in health. 2008;11(1):44–7.

28. Discacciati A, Bellavia A, Lee JJ, Mazumdar M, Valeri L. Med4way: a Stata command to investigate mediating and interactive mechanisms using the four-way effect decomposition. Oxford University Press; 2019.

29. Bland JM, Altman DG. Multiple significance tests: the Bonferroni method. BMJ. 1995;310(6973):170.

30. Finnikin S, Ryan R, Marshall T. Cohort study investigating the relationship between cholesterol, cardiovascular risk score and the prescribing of statins in UK primary care: study protocol. BMJ Open. 2016;6(11).

31. Pate A, Emsley R, Van Staa T. Impact of lowering the risk threshold for statin treatment on statin prescribing: a descriptive study in English primary care. British Journal of General Practice. 2020.

32. Martin A, Saunders CL, Harte E, Griffin SJ, MacLure C, Mant J, Meads C, Walter FM, Usher-Smith JA. Delivery and impact of the NHS health check in the first 8 years: a systematic review. British Journal of General Practice. 2018;68(672):e449–e59.

33. Deshpande S, Quek RG, Forbes CA, de Kock S, Kleijnen J, Gandra SR, Simpson Jr RJ. A systematic review to assess adherence and persistence with statins. Current Medical Research and Opinion. 2017;33(4):769–78.

34. Ingersgaard MV, Helms Andersen T, Norgaard O, Grabowski D, Olesen K. Reasons for nonadherence to statins–a systematic review of reviews. Patient preference and adherence. 2020:675–91.

35. Khatib R, Marshall K, Silcock J, Forrest C, Hall AS. Adherence to coronary artery disease secondary prevention medicines: exploring modifiable barriers. Open Heart. 2019;6(2):e000997.

36. Marcum ZA, Gellad WF. Medication adherence to multi-drug regimens. Clinics in geriatric medicine. 2012;28(2):287.

37. Grant RW, Devita NG, Singer DE, Meigs JB. Polypharmacy and medication adherence in patients with type 2 diabetes. Diabetes care. 2003;26(5):1408–12.

38. Grant RW, O’Leary KM, Weilburg JB, Singer DE, Meigs JB. Impact of concurrent medication use on statin adherence and refill persistence. Archives of internal medicine. 2004;164(21):2343–8.

39. Chodick G, Shalev V, Gerber Y, Heymann AD, Silber H, Simah V, Kokia E. Long-term persistence with statin treatment in a not-for-profit health maintenance organization: a population-based retrospective cohort study in Israel. Clinical therapeutics. 2008;30(11):2167–79.

40. Morotti K, Lopez J, Vaupel V, Swislocki A, Siegel D. Adherence to and persistence with statin therapy in a veteran population. Annals of Pharmacotherapy. 2019;53(1):43–9.

41. Svensson E, Nielsen RB, Hasvold P, Aarskog P, Thomsen RW. Statin prescription patterns, adherence, and attainment of cholesterol treatment goals in routine clinical care: a Danish population-based study. Clinical epidemiology. 2015:213–23.

42. NHS England. Quality and Outcomes Framework: NHSE; 2025 [11/04/2025]. Available from: https://qof.digital.nhs.uk/.

43. Brookhart MA, Patrick AR, Schneeweiss S, Avorn J, Dormuth C, Shrank W, Van Wijk BL, Cadarette SM, Canning CF, Solomon DH. Physician follow-up and provider continuity are associated with long-term medication adherence: a study of the dynamics of statin use. Archives of internal medicine. 2007;167(8):847–52.

44. Chan DC, Shrank WH, Cutler D, Jan S, Fischer MA, Liu J, Avorn J, Solomon D, Brookhart MA, Choudhry NK. Patient, physician, and payment predictors of statin adherence. Medical care. 2010;48(3):196–202.

45. Kampouraki E. 210 A positive consultation experience is associated with better statin adherence. BMJ Publishing Group Ltd and British Cardiovascular Society; 2023.

